# Comparative Study of Arteriosclerosis in Young and Elderly Individuals Using Carotid Artery Ultrasonography

**DOI:** 10.1101/2023.11.23.23298973

**Authors:** Ikuko Kawagishi, Kenichi Isobe, Takashi Koshikawa, Masaya Murase, Yoshiharu Naiki, Kosei Ota, Yosuke Ohira, Haruka Kondo, Takumi Shinohara, Yuto Shimazaki, Kakeru Takagi, Ayaka Takahashi, Marina Yamamoto, Atsushi Yoshida, Yoshitaka Ando, Kazunori Ohnishi

**Author notes:** Corresponding author: Kazunori Ohnishi, M.D., Department of Clinical Laboratory, Faculty of Medical Science, Shubun University, Nikko-cho 6, Ichinomiya, Aichi, 491-0938, Japan.

## Abstract

**Background:** Atherosclerosis-related strokes in middle-aged to elderly individuals have become a significant concern due to the risk to their lives and the deterioration of their quality of life. Ultrasonographic evaluation is becoming evident that it is useful for preventing strokes. However, many studies to date have used middle-aged and older subjects.

**Objective:** Since Shubun University has many students, as well as older faculty and staff, we searched for how effective carotid artery ultrasonographic evaluation is in predicting future cerebral infarction and other vascular disorders by comparing arterial stiffness between young and elderly subjects.

**Methods:** We examined the carotid arteries of 89 healthy adults (37 males and 52 females) from Shubun University, ranging in age from 19 to 77 years old. We measured the intima-media thickness (IMT), the plaques, and the vessel diameter following the guidelines for carotid artery ultrasonography.

**Results:** IMT showed a significant age-related increase in both men and women in various parts of the carotid artery, except for women in the bifurcation area in our study. The overall max IMT of the carotid artery showed a significant age-related increase in both men and women. While carotid artery IMT did not change much until the 40s, there was a trend of thickening in the 50s to 60s. Plaque formation began to appear around the 40s. Age-related changes in vascular diameter were more pronounced in older men, particularly those aged 50 and above.

**Conclusions:** The results revealed that IMT, the number of plaques and the inner diameter of the carotid artery, remained largely unchanged up to the age of 40, after which they began to increase.

## Introduction

Cerebral infarction caused by arteriosclerosis in middle-aged to elderly people has become a major problem in elderly people due to the danger to their lives and the deterioration of their quality of life, and the possibility of predicting and preventing atherosclerosis has become a hot topic worldwide.^1,2^ X-ray diagnostics, represented by CT, are widely used for the preventive diagnosis of atherosclerosis. However, the use of X-rays and contrast agents can be harmful to the body. Additionally, the equipment required for X-ray diagnosis is large and expensive. Ultrasonographic evaluation, on the other hand, do not harm the body, are relatively cost-effective, and the equipment is compact and easily portable. In recent years, their use at the bedside has also increased. Carotid artery ultrasonography, including in Japan, has been widely used worldwide, and its usefulness has become apparent.^3,4^

Carotid artery ultrasonography is often used to measure 1) intima-media thickness (IMT), 2) the number of plaques, and 3) the inner diameter of the carotid artery as standard parameters. In Japan, in 2017, the the Japan Society of Ultrasonics in Medicine and the Japan Academy of Neurosonology jointly created guidelines for the standard evaluation of carotid artery lesions using ultrasonography, leading to more objective indicators.^5^ As for age-related carotid artery changes incorporated in the guidelines, intima-media complex thickness (IMT), which is the combined thickness of the intima and tunica media, increases relatively linearly at 0.009 mm/year in healthy adults.^6^ However, these data are based on healthy adults aged 30 and above, and a comparison with younger generations is warranted. Since Shubun University has many students, as well as older faculty and staff, we began this study to compare arterial sclerosis between younger and older adults to search for how effective cervical artery ultrasonography is in predicting future cerebral infarction and other vascular disorders. We investigated the following parameters: intima-media thickness (IMT), which is approximately 0.6 mm at the age of 30 and is reported to increase by 0.1 mm every 10 years, to determine whether this holds true. We also examined when plaques tend to appear and whether there are plaques at the right subclavian artery branch. Additionally, we explored whether vascular diameter expands with age.

## Methods

### Subjects

In this study, we examined the carotid arteries of 89 healthy adults and assessed the measured values by age and gender. The participants included a total of 89 individuals (37 males and 52 females) from Shubun University, ranging in age from 19 to 77 years old, with a mean age of 33.7 years (42.5 years for males and 27.4 years for females). The median age was 21 years old (39 males and 21 females) (Table 1). All students were in their 20s or younger. The faculty consisted of one man and one woman in their 20s, while the others were aged 30s or older. This study was conducted with the approval of the Research Ethics Committee of Shubun University Faculty of Medical Science (R05-0001).

**Table 1.** Age distribution of subjects whose cervical arteries were measured.

|  | 10s | 20s | 30s | 40s | 50s | 60s | 70s |
| --- | --- | --- | --- | --- | --- | --- | --- |
| Male (n=37) | 0 | 16 | 3 | 2 | 3 | 8 | 5 |
| Female (n=52) | 2 | 36 | 6 | 2 | 4 | 8 | 0 |

\* All students were in their 20s or younger. The faculty consisted of one man and one woman in their 20s, while the others were aged 30s or older.

### Ultrasonographic device and probe selection

The device used in this study was Sefius UF-890AG (the Fukuda Electronic Co., Ltd.), and we utilized a linear probe with a central frequency of 5-12 MHz.

### Carotid artery ultrasonographic imaging procedure

Patients were positioned supine without the use of a pillow. On the right side, the common carotid artery was observed until it was no longer visible toward the subclavian artery bifurcation. On the left side, the aortic bifurcation was followed until it was no longer visible. The patient was then asked to face diagonally up to the left and the probe was placed in the middle of the neck and observed in the short axis toward the head. Subsequently, observation was made in the long axis. Observation was made from the common carotid artery to the bifurcation, the internal carotid artery, and the point where the external carotid artery was no longer visible.

### Measurement points and methods

Following the guidelines for carotid artery ultrasonography,^5^ we measured 1) intima-media thickness (IMT), 2) plaques, and 3) vessel diameter. IMT measurements were taken at the thickest IMT point in the common carotid artery, IMT at C10 (C10 refers to the area 10 mm from the origin of the bifurcation toward the common carotid artery, and the mean IMT was defined as the IMT at this area), and the thickest IMT at the branching point and internal carotid artery. Vessel diameter was measured at C10 in the common carotid artery, and at the portion of the internal carotid artery and vertebral artery where the diameter was kept constant. Plaques were observed in the subclavian bifurcation as well as in the carotid artery as raised lesions larger than 1.1 mm, and were measured if present. Statistical analysis was performed using the statistical software EZR R4.3.1, with Spearman’s rank correlation coefficient for correlation with age, Mann-Whitney U test for comparison between students and faculty, and Wilcoxon rank sum test for right-left difference in vessel diameter, respectively.

## Results

### 1. Age-related changes in carotid artery IMT

(1) Age-related changes in common carotid artery IMT at C10 Age-related changes in IMT of the common carotid artery were measured in the right (R) and left (L) C10 regions in men and women. The age-related changes in C10 IMT were statistically significant in both the right and left sides for males (right: p=0.0017, left: p<0.0001), and in the left side for females (right: p=0.48, left: p=0.04). C10 IMT showed an increasing trend in both males and females, with a noticeable increase occurring around the age of 50 (Figures 1-1,3). In males, there were a higher number of participants in their 20s, leading to smaller IMT values compared to other age groups. IMT values in the 30s to 50s showed little change. However, after the age of 50, IMT values clearly increased proportionally with age. Age-specific averages by decade are shown in Figure 1-2. Looking at the averages, IMT in the 20s was 0.43 mm (R) and 0.41 (L), IMTs in the 30s were 0.43 (R) and 0.4 (L) with little variation, and in the 40s, they decreased slightly to 0.35 (R) and 0.3 (L). However, the sample sizes were small in the 30s and 40s, so these changes are likely due to individual characteristics. In the 50s, IMTs increased to 0.53 (R) and 0.47 (L), and this trend continued in the 60s with 0.56 (R) and 0.66 (L). In females, IMT showed no significant changes until their 40s. However, similar to males, after the age of 50, an increasing trend was observed in the 50s [0.4 (R) and 0.5 (L)] and 60s [0.6 (R) and 0.65 (L)] (Figures 1-2,4).

**Figure 1-1.**
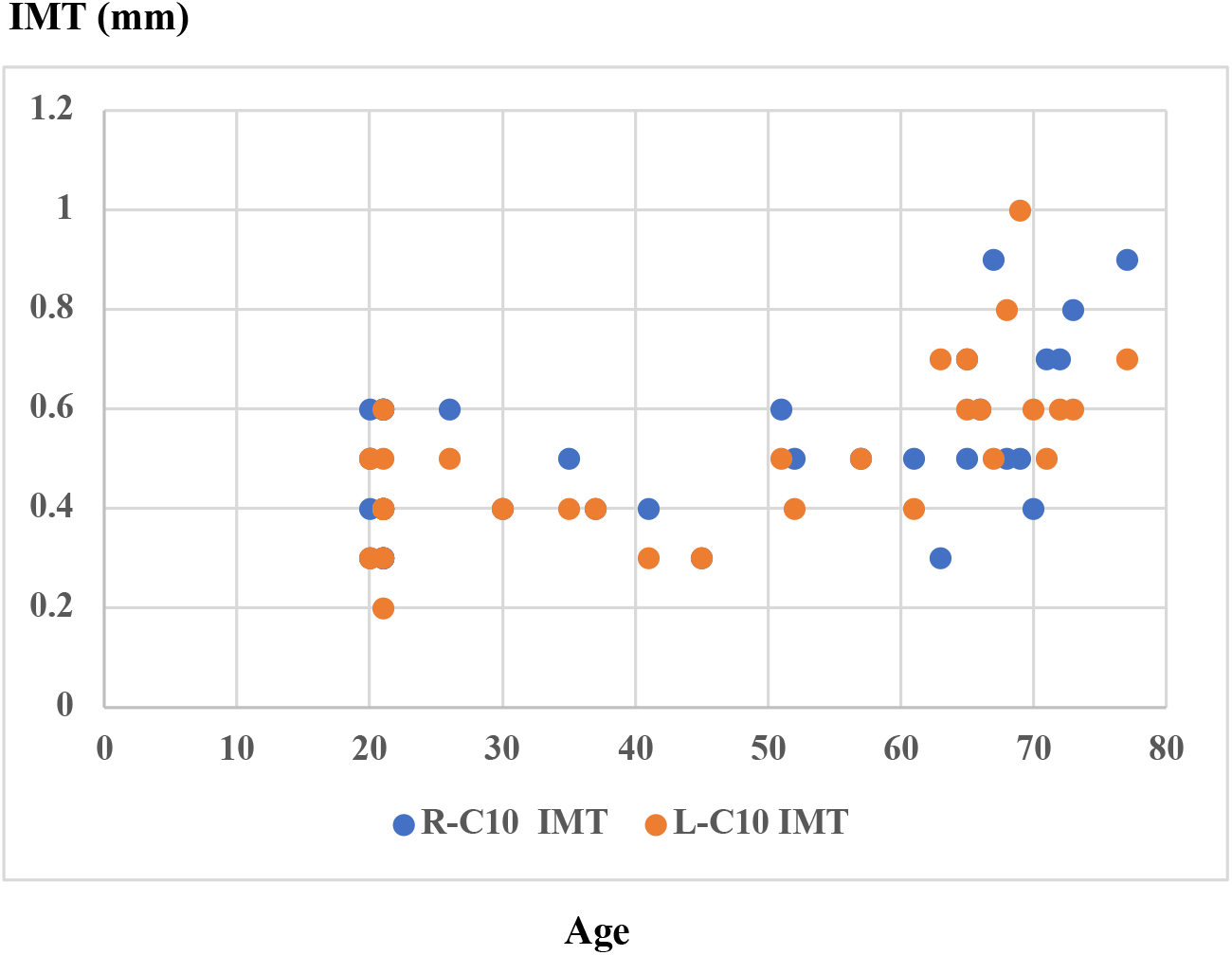
The age-related changes in C10 IMT (male) *Spearman’s rank correlation coefficient, right: p=0.0017, left: p<0.0001 **Abbreviation: IMT, intima-media thickness *** C10 refers to the area 10 mm from the origin of the bifurcation toward the common carotid artery

**Figure 1-2.**
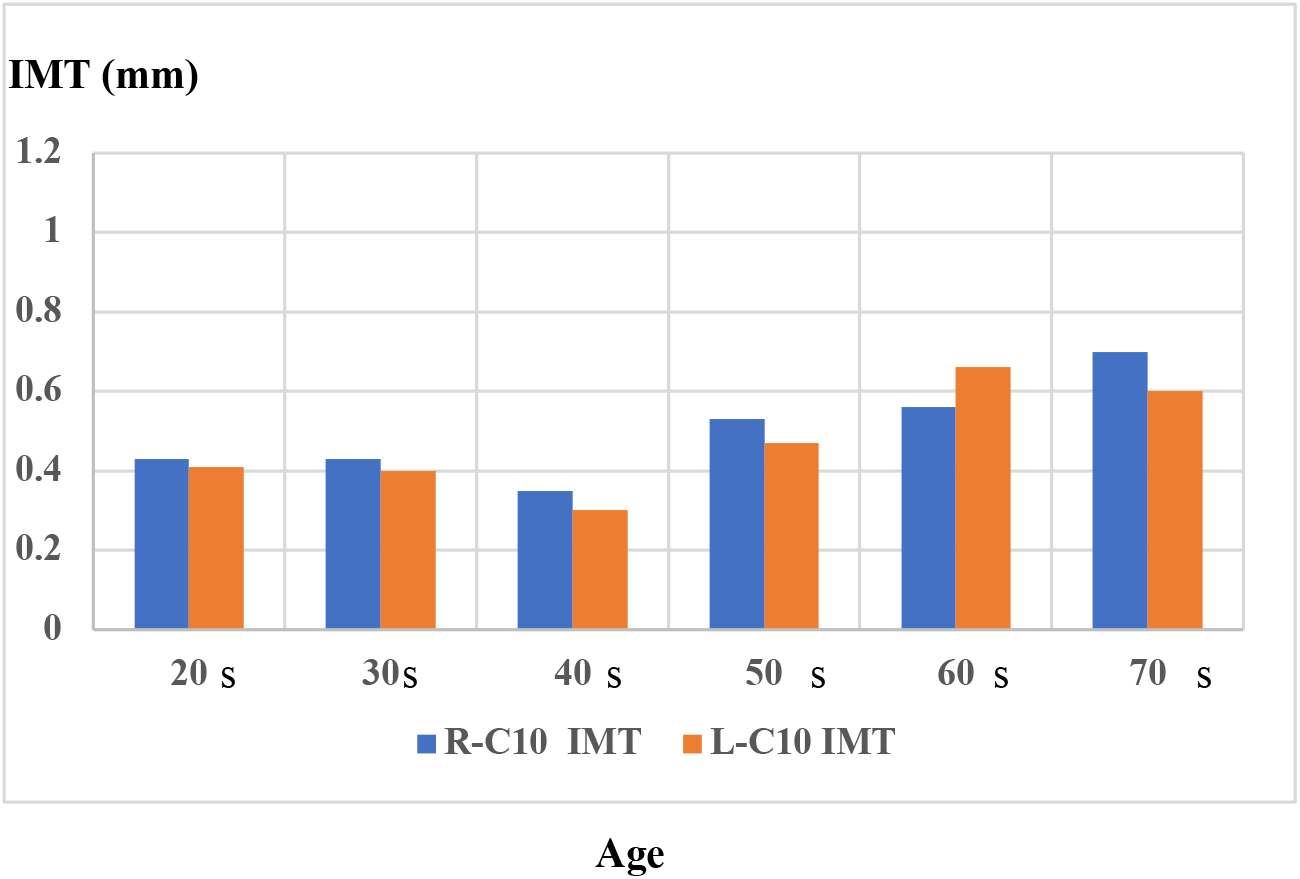
Age-specific average values of C10 IMT (male)

**Figure 1-3.**
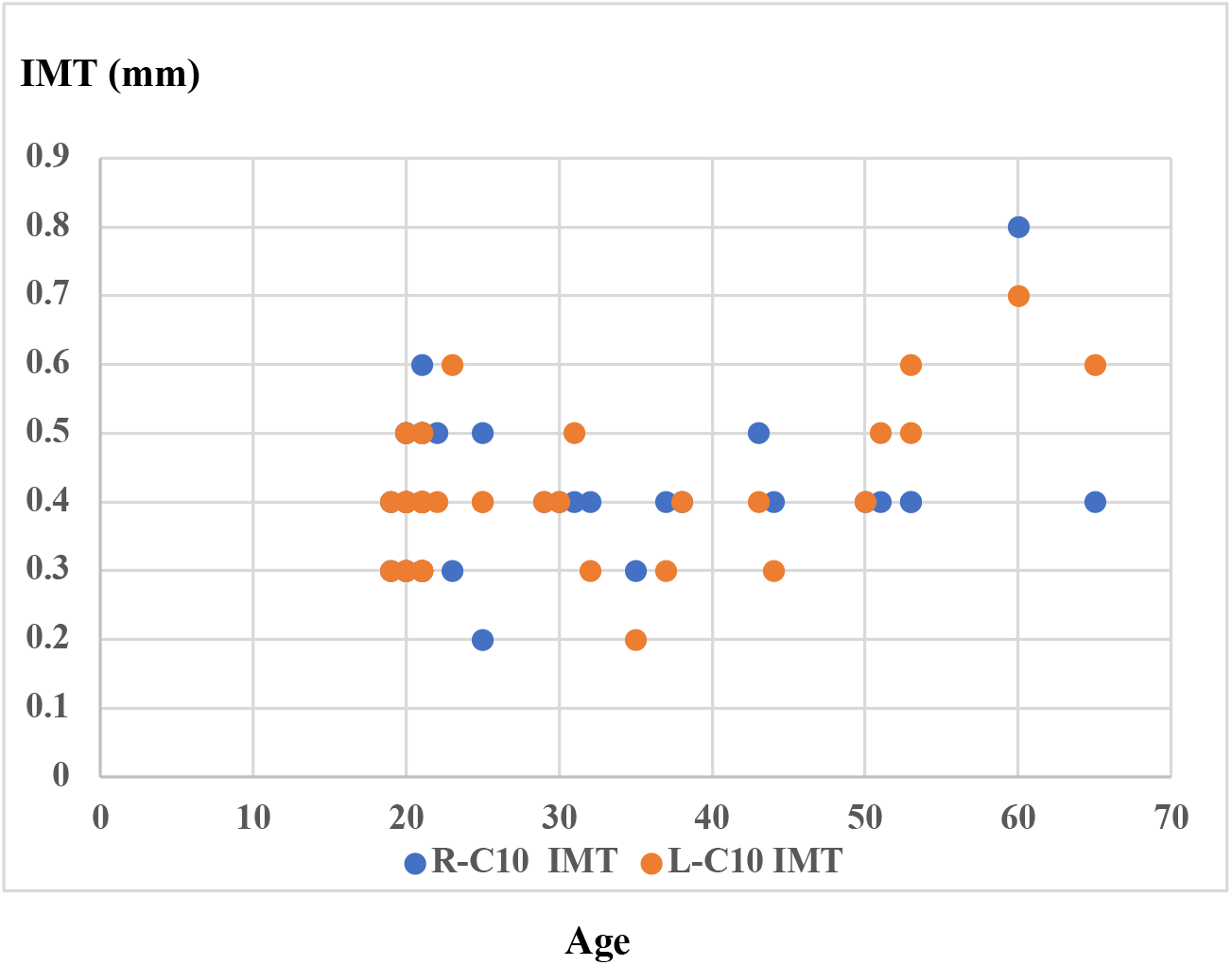
The age-related changes of C10 IMT (female) *Spearman’s rank correlation coefficient, right: p=0.48, left: p=0.04

**Figure 1-4.**
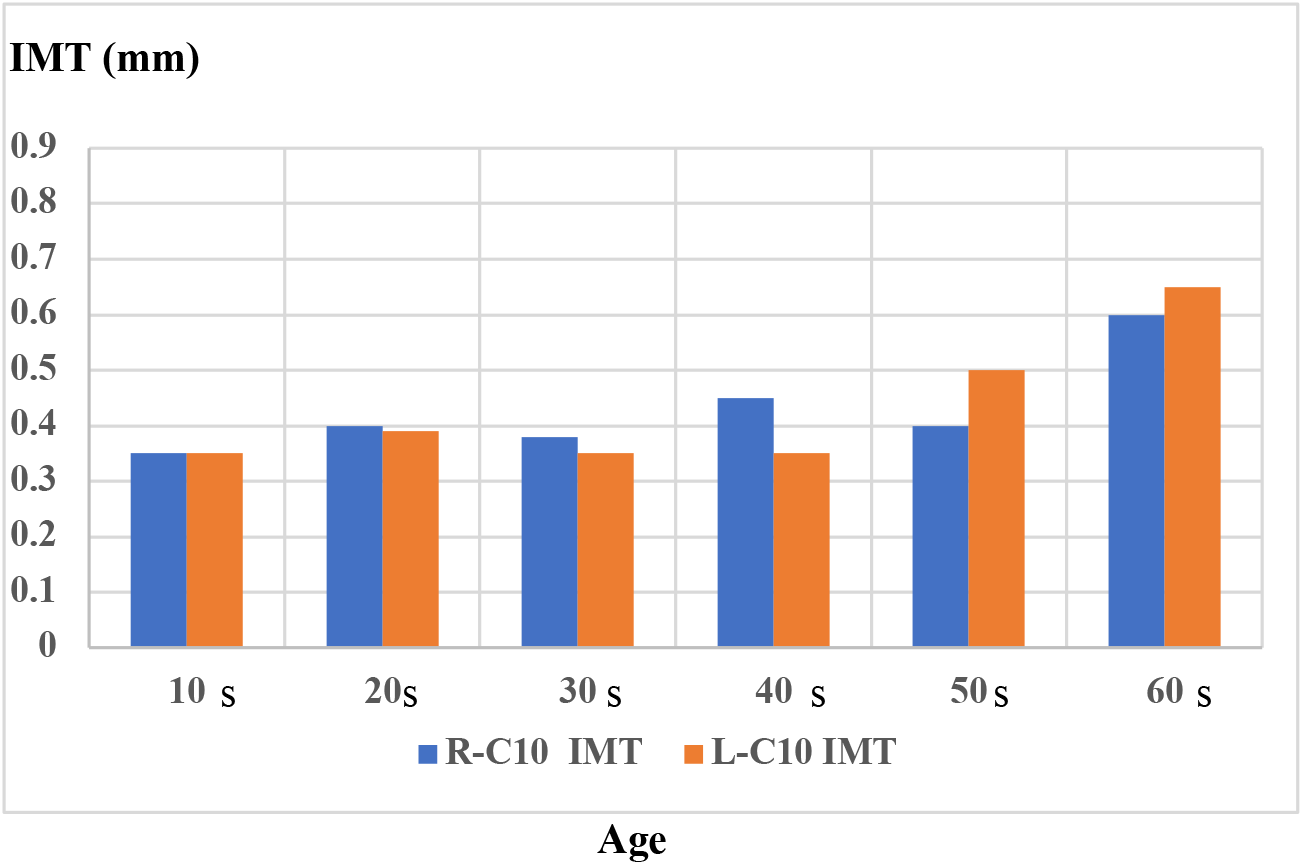
Age-specific average values of C10 IMT (female)
(2) Age-related changes in common carotid artery (CCA) max IMT The age-related changes in CCA max IMT were similar to those of C10 IMT. In males, CCA max IMT significantly increased in both the right and left sides (right: p=0.0003, left: p<0.0001), and in females, it increased significantly in the left side (right: p=0.337, left: p=0.034). CCA max showed an increasing trend in both males and females, with a noticeable increase occurring around the age of 50 (Figures 2-1,2).

**Figure 2-1.**
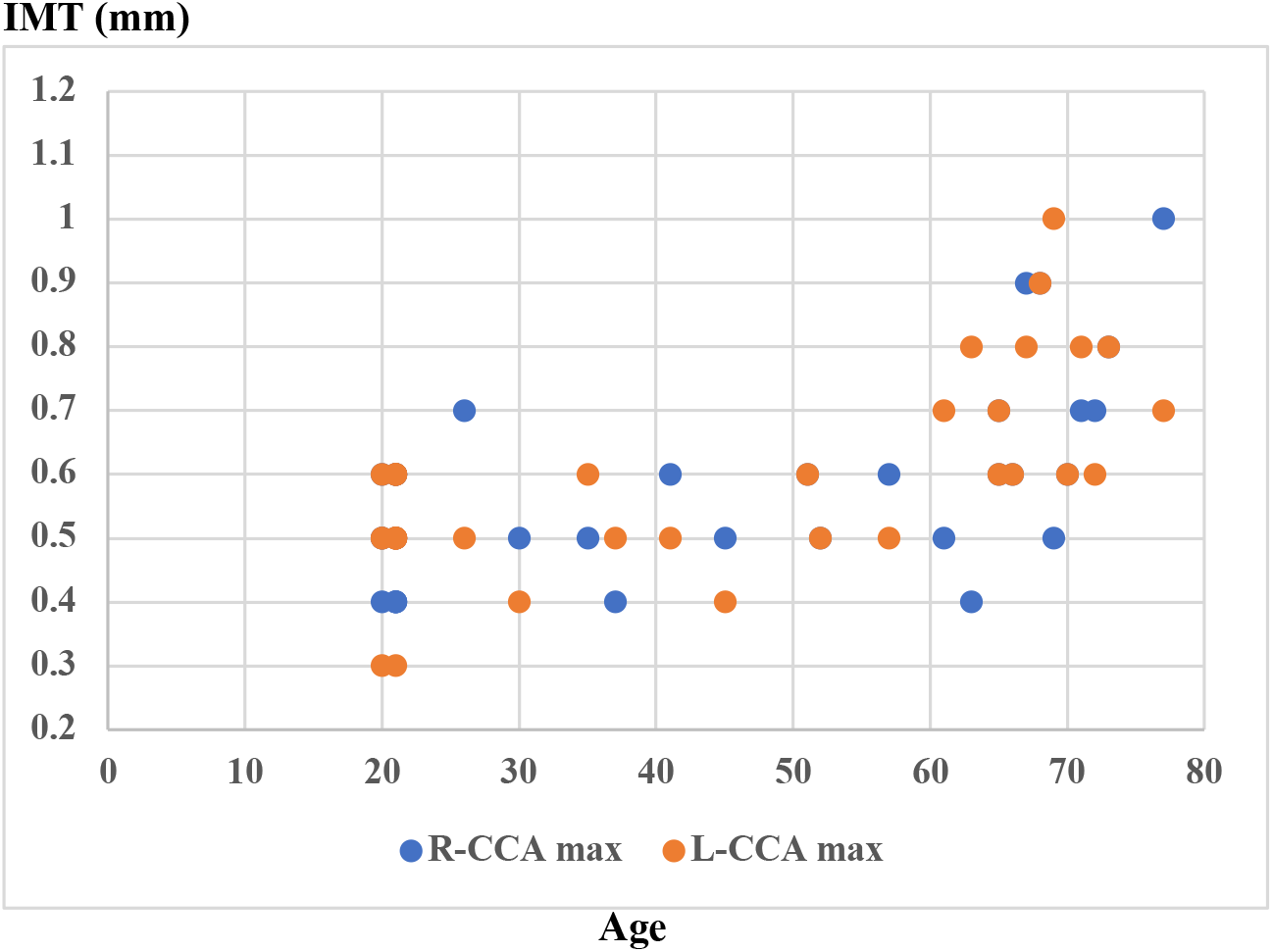
Age-related changes in CCA max IMT (male) *Spearman’s rank correlation coefficient, right: p=0.0003, left: p<0.0001 **Abbreviation: CCA, common carotid artery

**Figure 2-2.**
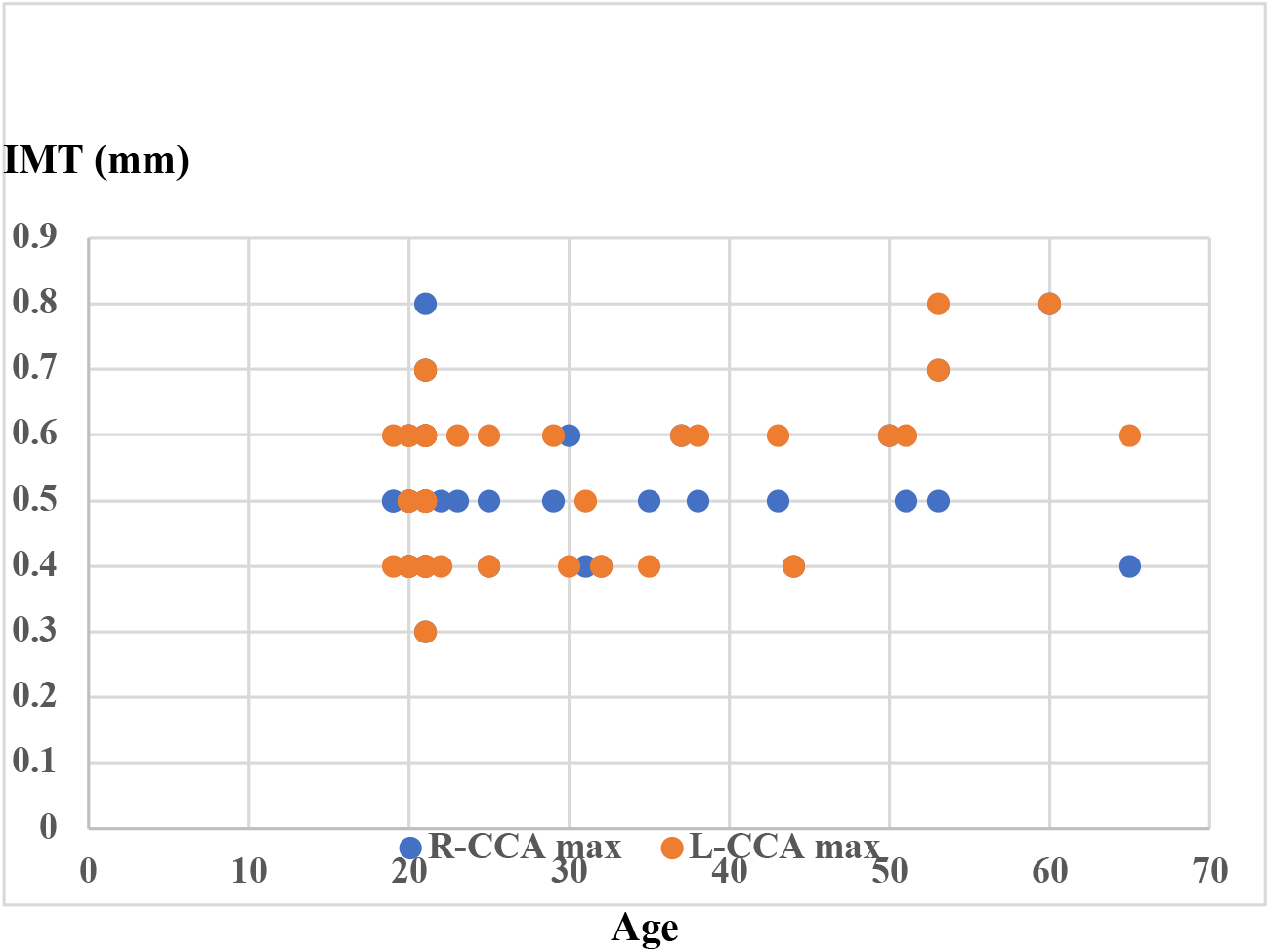
Age-specific average values of CCA max IMT (female) *Spearman’s rank correlation coefficient, right: p=0.337, left: p=0.034
(3) Age-related changes in bifurcation (Bif) max IMT Bif max IMT showed statistically significant age-related increases in both males and females (males, right: p<0.0001, left: p<0.0001; females, right: p=0.022, left: p<0.0015). Particularly, in males, there was a noticeable increase in the 50s and above, while in females, the increase trend was more pronounced in the 60s and above (Figures 3-1,2).

**Figure 3-1.**
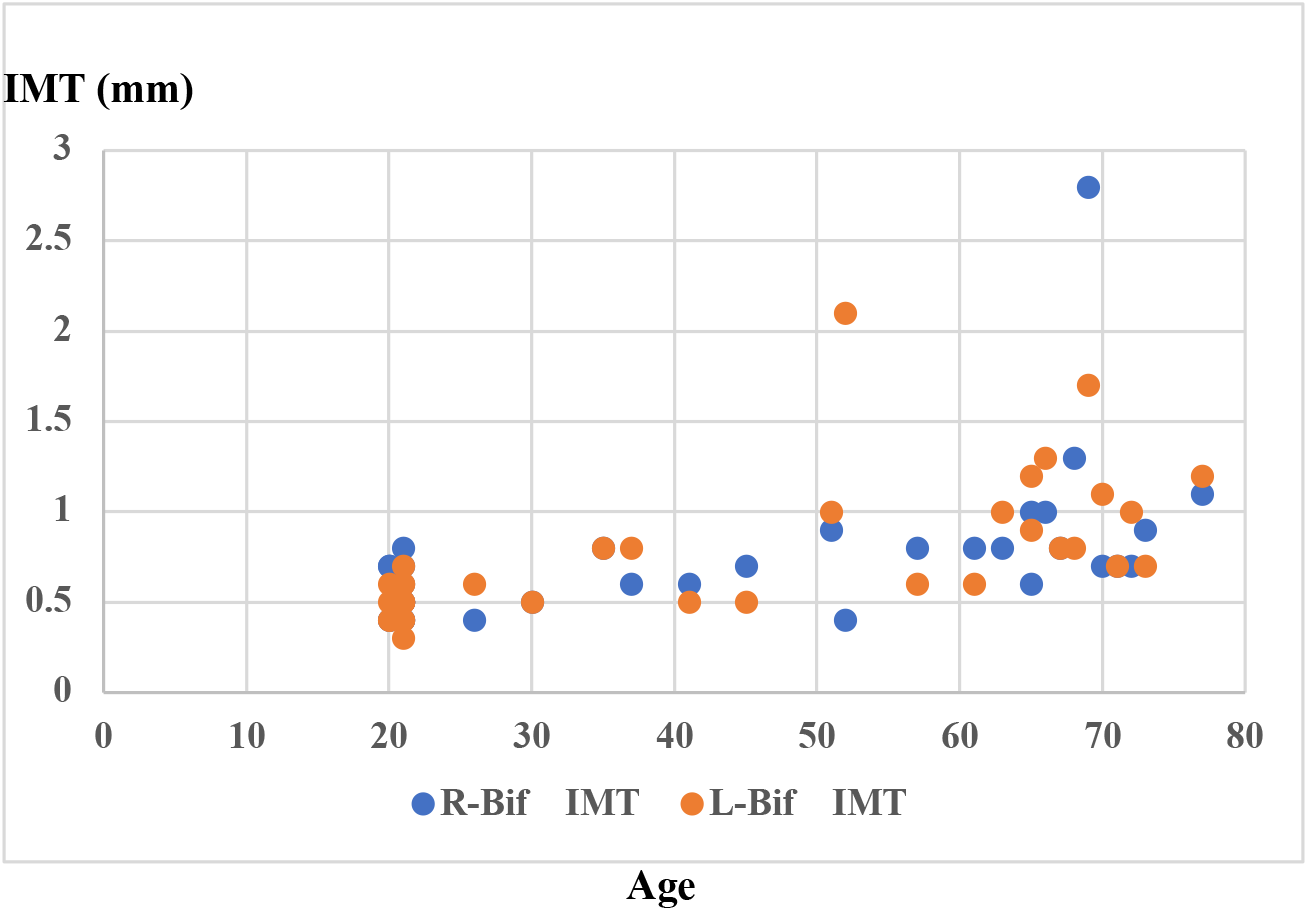
Age-related changes in Bif max IMT (male) *Spearman’s rank correlation coefficient, right: p<0.0001, left: p<0.0001 **Abbreviation: Bif, bifurcation

**Figure 3-2.**
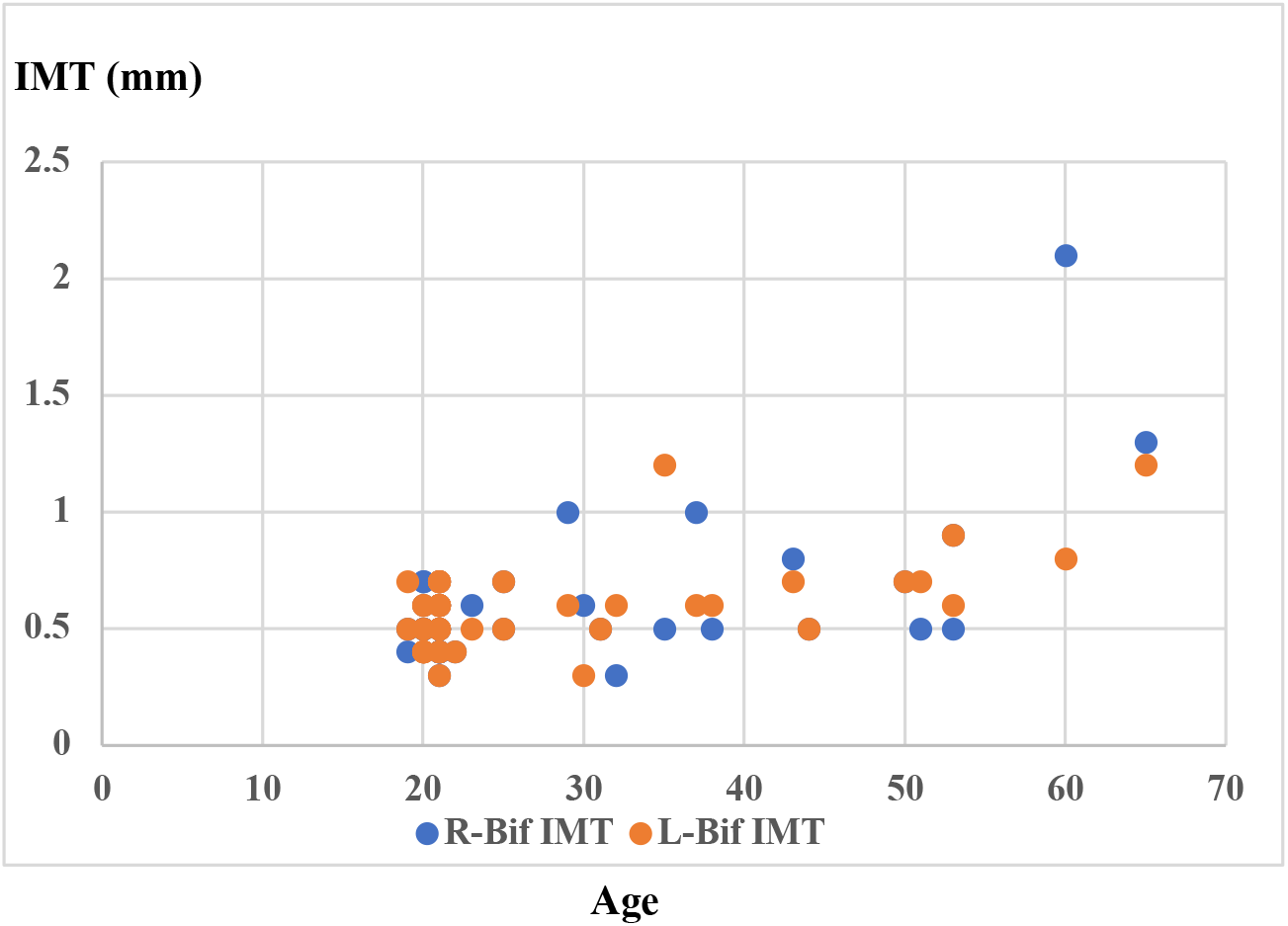
Age-related changes in Bif max IMT (female) *Spearman’s rank correlation coefficient, right: p=0.022, left: p<0.0015
(4) Age-related changes in internal carotid artery (ICA) max IMT The age-related changes in ICA max IMT showed statistically significant increases in both the right and left sides for males (right: p=0.0072, left: p=0.0094) and in the left side for females (right: p=0.90, left: p=0.006) (Figures 4-1,2).

**Figure 4-1.**
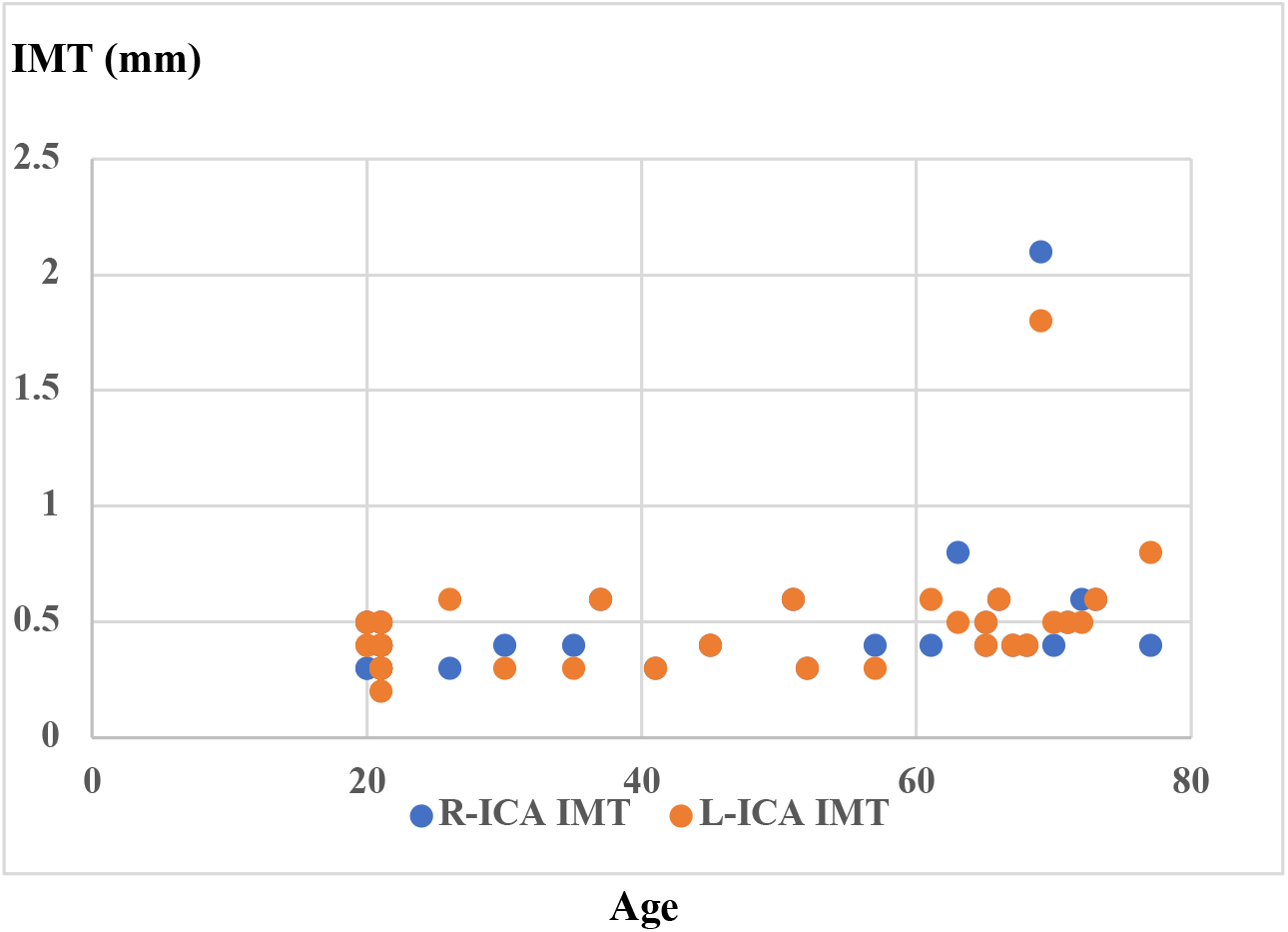
Age-related changes in ICA max IMT (male) *Spearman’s rank correlation coefficient, right: p=0.0072, left: p=0.0094 ** Abbreviation: ICA, internal carotid artery

**Figure 4-2.**
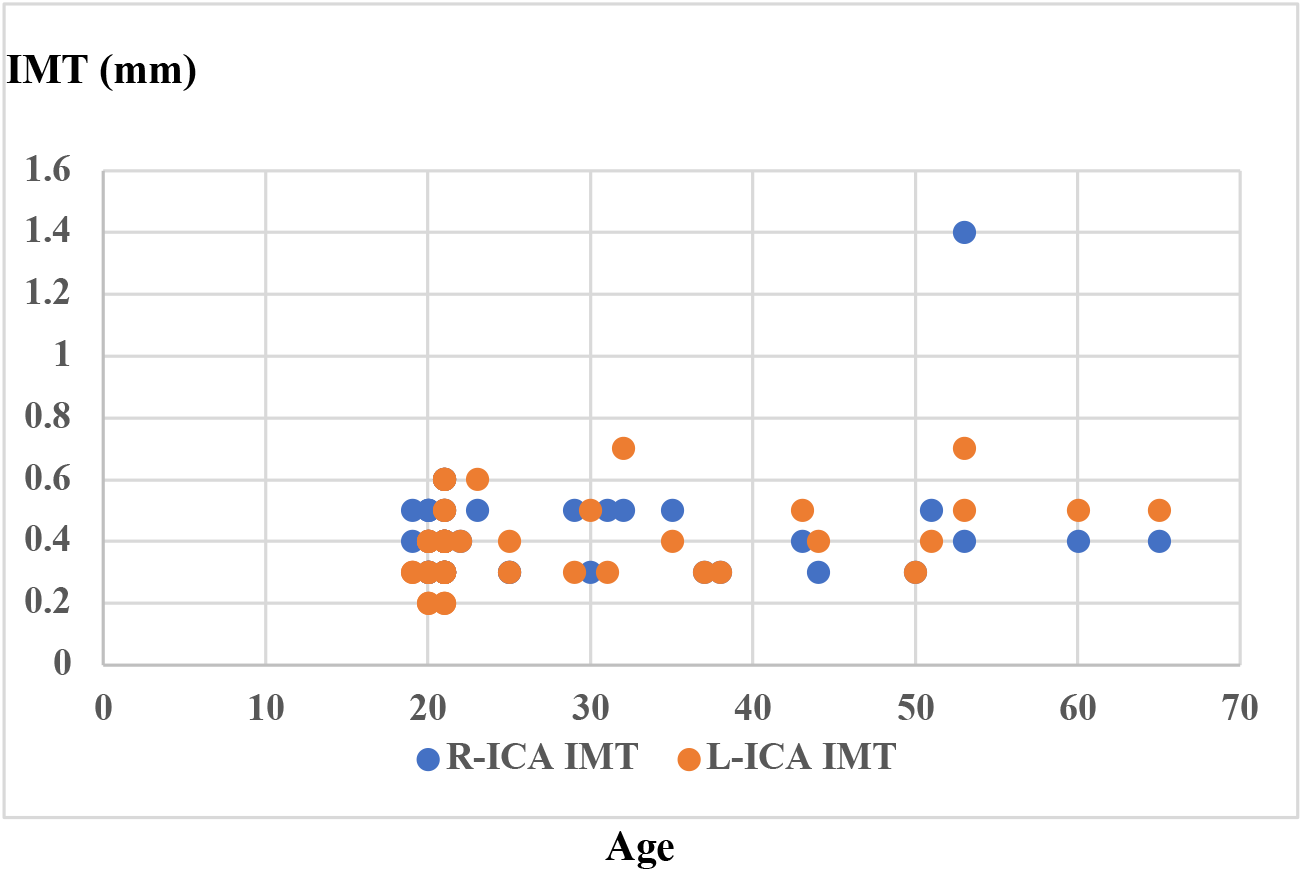
Age-related changes in ICA max IMT (female) *Spearman’s rank correlation coefficient, right: p=0.90, left: p=0.006
(5) Age-related changes in max IMT across the entire carotid artery Max IMT across the entire carotid artery was measured by taking the thickest max IMT in both the right and left sides for males and females. Max IMT showed statistically significant age-related increases in both males (p<0.0001) and females (p<0.0019). Particularly, max IMT tended to increase in men in their 50s and older, and in women in their 60s and older (Figures 5-1,2).

**Figure 5-1.**
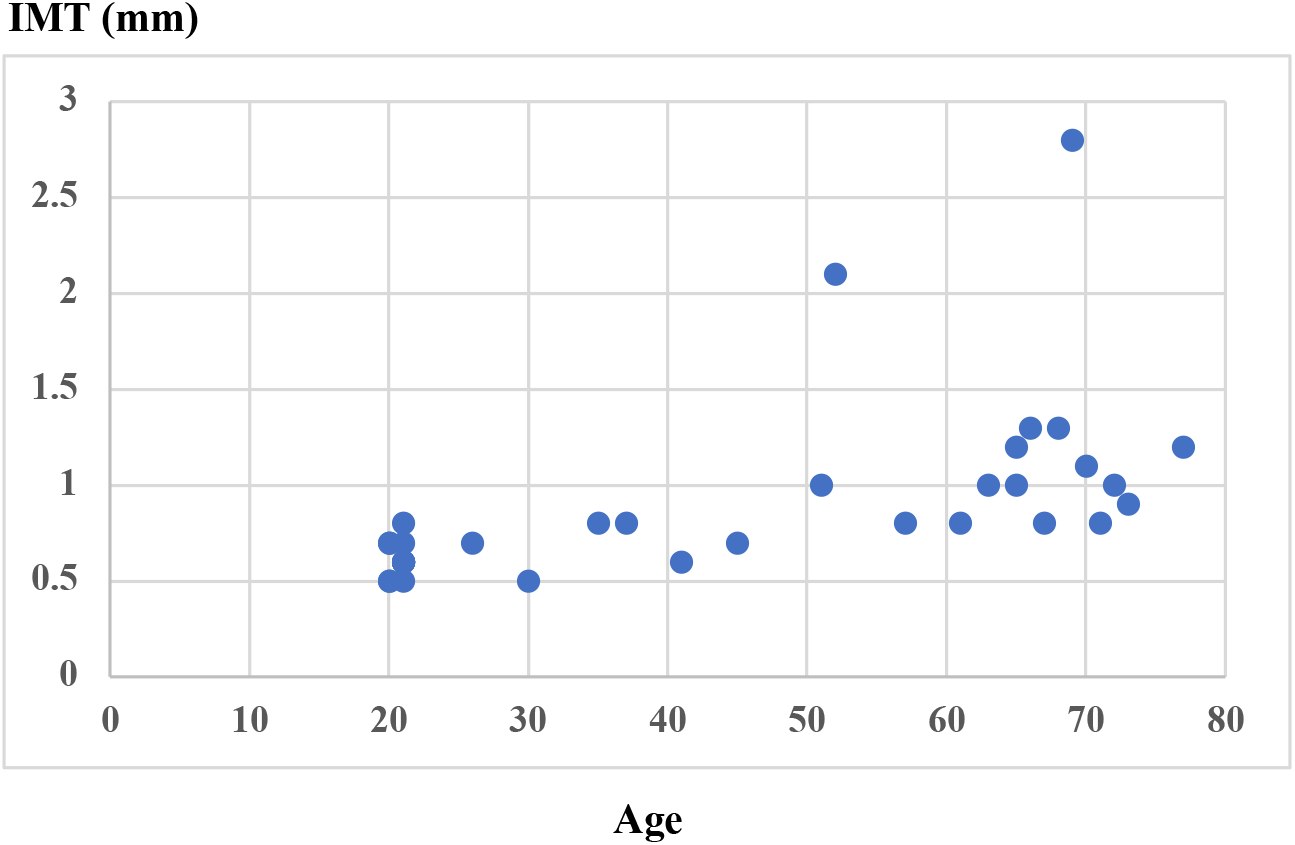
Age-related changes in max IMT in the entire carotid artery (male) *Spearman’s rank correlation coefficient, p<0.0001

**Figure 5-2.**
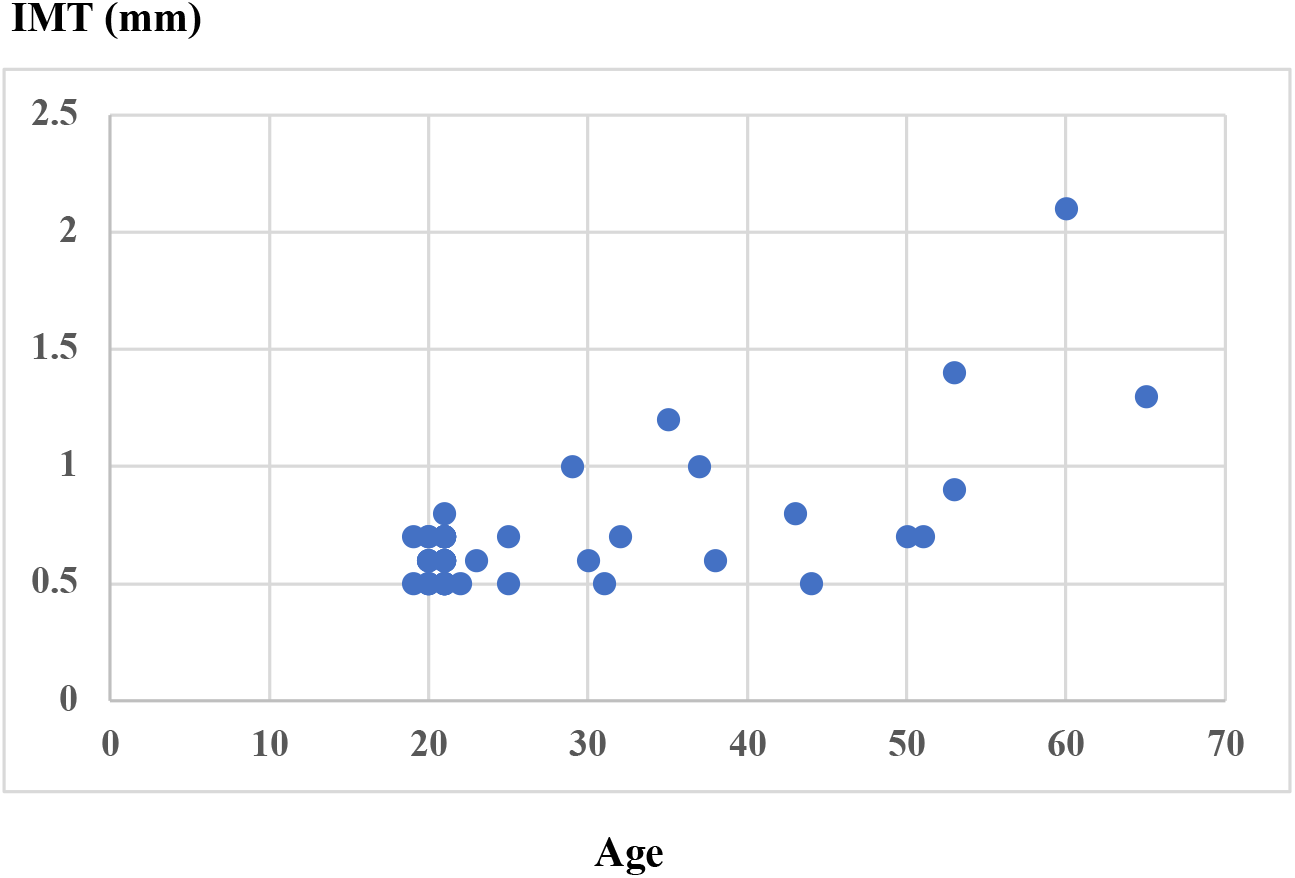
Age-related changes in max IMT in the entire carotid artery (female) *Spearman’s rank correlation coefficient, p<0.0019
(6) Comparison of IMT including plaque between students and faculty at all sites The IMT of each site was significantly increased in both the right and left sides for all sites for faculty compared to students (p<0.02). Specifically, faculty members exhibited higher IMT values in both the right and left sides at all measurement sites. Notably, when comparing male faculty aged 50 and above with male students, significant increases were observed in both the right and left sides at all measurement sites (Figure 6). In females, only the bifurcation (Bif) site showed significantly higher IMT in faculty compared to students (p<0.02), while there were no significant differences in other measurement sites. Comparisons could not be made for females because of the small number of faculty members over 50 years of age.

### 2. Frequency of plaque occurrence

Plaques were counted as localized lesions with IMT of 1.1 mm or greater. First, the occurrence of plaques in the right subclavian artery branching point and the common carotid artery was counted by gender and age. In the subclavian branching point, plaques were observed in nine individuals aged 40 and above, while in the common carotid artery, they appeared in seven individuals aged 60 and above. For both males and females, plaques were sporadically observed up to the age of 60, but they became significantly more frequent in individuals aged over 60 (Table 2).

**Figure 6.**
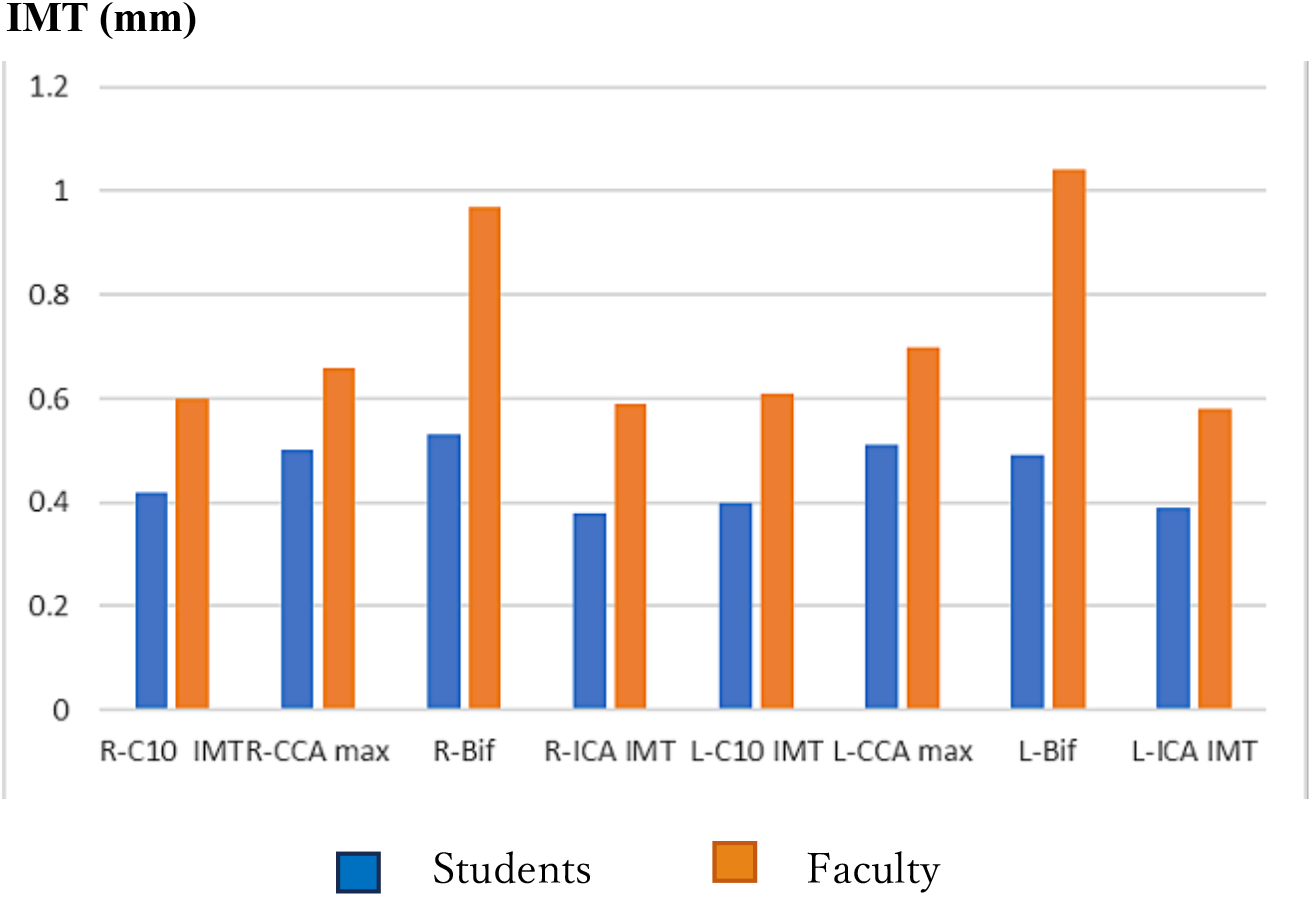
Comparison of mean IMT between students and faculty aged 50 and above at all sites (male) *Mann-Whitney U test, p<0.02 in both the right and left sides for all sites

**Table 2.** Subjects with plaque in R-SCA or carotid artery.

|  | R-SCA |  | Carotid artery |  |
| --- | --- | --- | --- | --- |
| Age | Male (n=37) | Female (n=52) | Male (n=37) | Female (n=52) |
| 10s |  | 0 |  | 0 |
| 20s | 0 | 0 | 0 | 0 |
| 30s | 0 | 0 | 0 | 1 |
| 40s | 1 | 0 | 0 | 0 |
| 50s | 1 | 1 | 0 | 0 |
| 60s | 4 | 1 | 3 | 2 |
| 70s | 1 |  | 2 |  |

\*\*Abbreviation: R-SCA, right subclavicular artery

### 3. Comparison of vascular diameter by gender and age

The age-related changes in vessel diameter were observed in C10, where both the right and left sides showed a significant increase in males (right: p<0.001, left: p=0.0019). However, no significant increase was observed in ICA (Figures 7-1,2).

**Figure 7-1.**
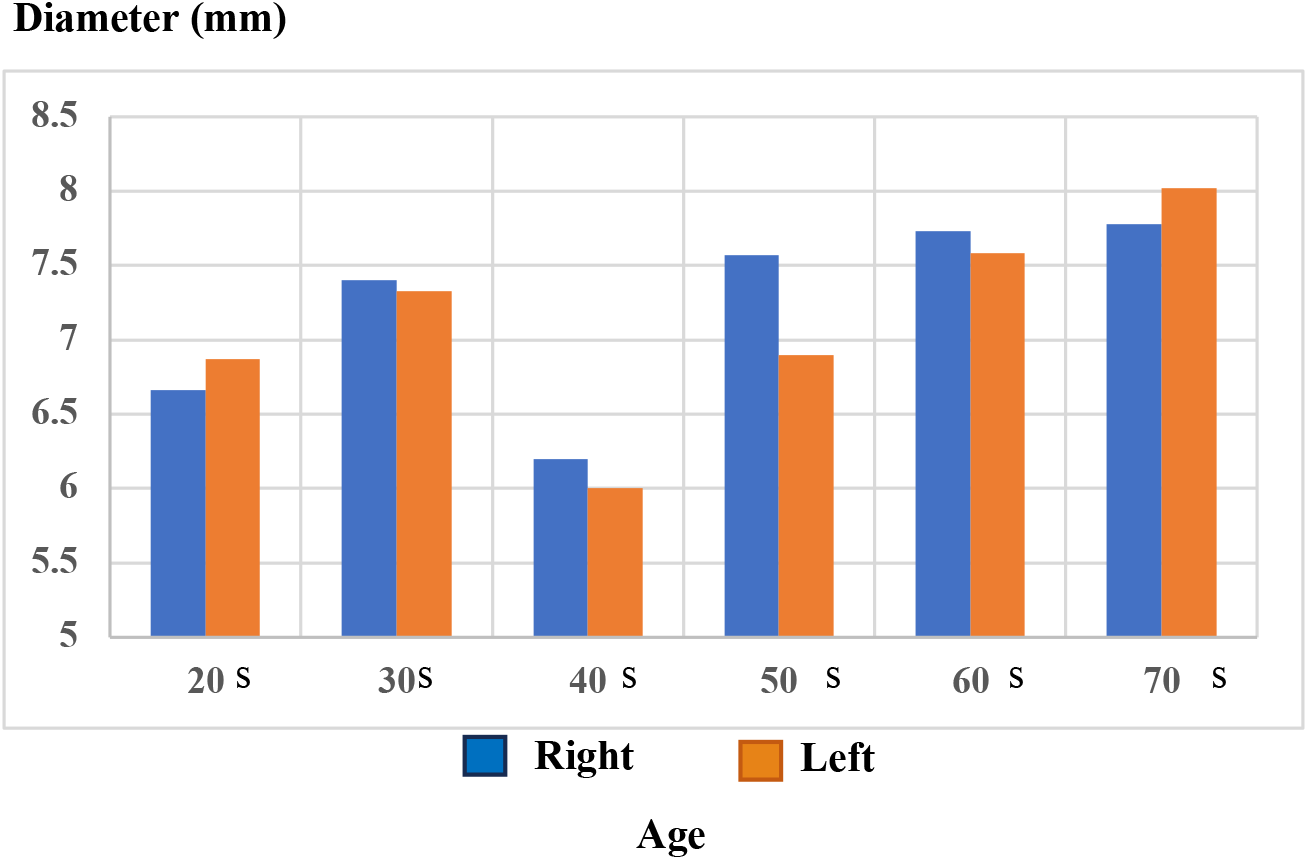
Age-related changes in mean C10 diameter (male) *Wilcoxon rank sum test, right: p<0.001, left: p=0.0019

**Figure 7-2.**
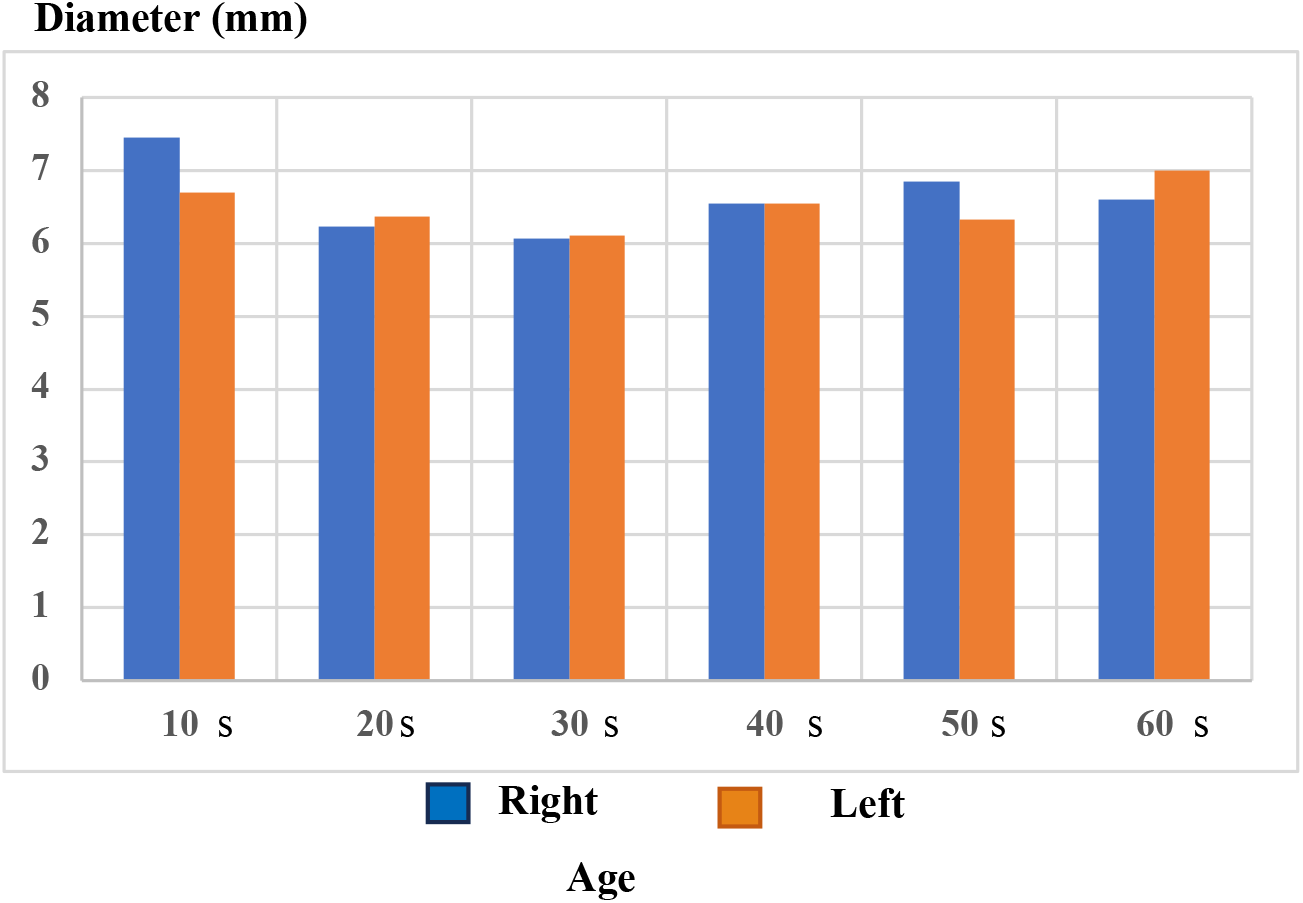
Age-specific changes in C10 diameter (female) *Wilcoxon rank sum test, not significant in both the right and left sides.

### 4. Comparison of vascular diameter between students and faculty

In the comparison of vascular diameter between students and faculty, in the C10 segment, both right and left sides showed a significant increase in vascular diameter among male faculty (right: p<0.001, left: p=0.014), while no significant difference was observed in female subjects. In the ICA segment, there were no significant differences in vascular diameter between males and females. Specifically, when comparing male faculty aged 50 and above with students, a significant increase in vascular diameter was observed in the C10 segment on both sides (p<0.001, p=0.004), while no significant differences were found in the ICA and the vertebral artery (VA) segments (Figure 8). However, there were too few female faculty members aged 50 and above to perform a meaningful comparison.

**Figure 8.**
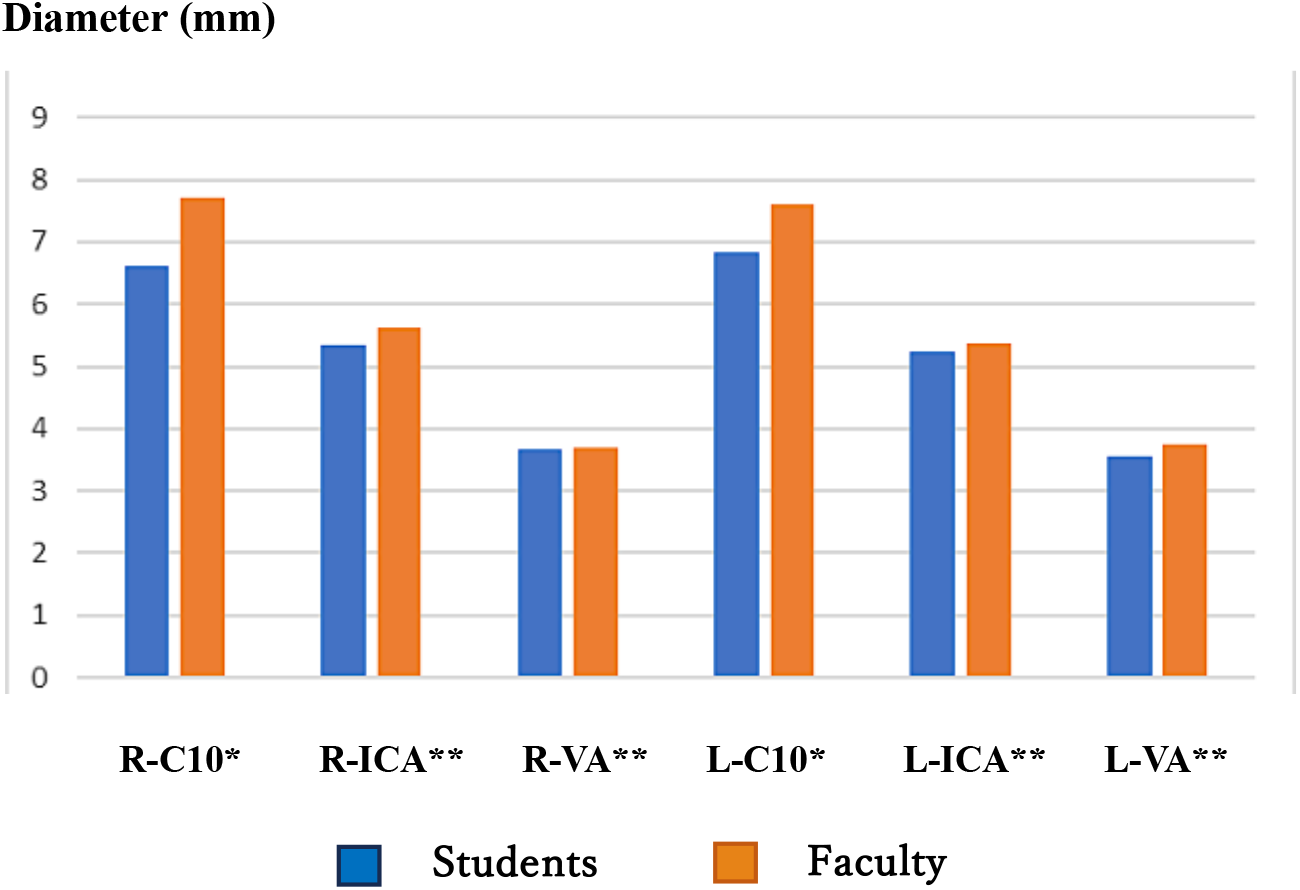
Comparison of mean vascular diameter between students and faculty aged 50 and above at all sites (male) *Mann-Whitney U test, right: p<0.001, left: p=0.014 ** Mann-Whitney U test, not significant ***Abbreviation: VA, vertebral artery

### 5. Lateral difference in vascular system

The right-left difference in vascular system was weakly significant only for females in VA (male: p=0.33, female: p=0.045).

## Discussion

Carotid echocardiography has gained popularity as a non-invasive and convenient diagnostic tool for the prevention of strokes associated with atherosclerosis. While many studies assessing its usefulness have focused on middle-aged and older individuals, this study aimed to compare a wider age range, from late teens to people in their 70s, to gain insights into younger individuals. The results of this study revealed that key indicators of atherosclerosis, such as intima-media thickness (IMT) and plaque count, tend to increase starting in the 50s when comparing younger individuals to this age group. This finding is consistent with the known increase in stroke incidence in this age range.

Here, we will first discuss whether carotid artery ultrasonography can serve as a predictive tool for strokes and even cardiovascular diseases such as myocardial infarction. Then, we will explore how the age-related changes in IMT, plaque, and vessel diameter measured by carotid artery ultrasonography compare to other reported findings.

### 1. Relationship between plaque, IMT, and atherosclerosis

Atherosclerotic plaques are initiated by factors such as high cholesterol, high blood pressure, smoking, and diabetes, which induce stress in the body. This stress triggers the oxidation of LDL (low-density lipoprotein), leading to its accumulation in the endothelium.^7,8^ The accumulation of oxidized LDL in the endothelium provokes inflammation, resulting in the formation of plaques characterized by lipid and smooth muscle cell debris encased within a fibrous cap. The rupture of these fibrous plaques is believed to lead to cardiovascular events, including myocardial infarction and stroke.^9^

### 2. Relationship between carotid IMT and disease

Since carotid intima media thickness (CIMT) directly reflects atherosclerosis in the region, its potential to cause occlusion of distant cerebral vessels can naturally be considered.^10^ Since carotid artery stiffness is a strong risk factor for stroke, CIMT is reported to be a simple and very good indicator.^11,12^ Furthermore, many reports have suggested that elevated CIMT is an indicator not only of cerebral infarction but also of other cardiovascular diseases.^13–20^ However, there are some papers that critically describe the relationship of CIMT to ischemic heart disease and cerebral infarction. One of the largest global studies to date is the Rotterdam study conducted in 1992-1993. In this study, 5,965 of 7,983 participants underwent carotid artery echocardiography. Of these, 140 had strokes and 125 had experienced myocardial infarctions. They concluded that CIMT is associated with stroke but not myocardial infarction.^21^ Hester et al. performed a meta-analysis from the Pubmed and Embase data base from 1950-2012 and concluded that CMIT is poorly associated with cardiovascular diseases.^22^ Thus, although there is a divergence of opinions regarding the association between CIMT and systemic cardiovascular system, there is no dispute that it is predictive of cerebral infarction. Based on these papers, studies on its prophylactic drug administration are being conducted both domestically and internationally.^23,24^

### 3. Comparison with other reports of CIMT with increasing age

Our CIMT measurement results, though from a limited sample with an age bias, showed that in the 20s, CIMT (C10 IMT) values for men were 0.43 (R) and 0.41 (L), while for women, they were 0.4 (R) and 0.39 (L). In the 30s, both men and women had slightly smaller values than these. In the 40s, there was little change from the values observed in the 20s. However, for those over 50, CIMT values increased significantly. For men in their 50s, CIMT was 0.53 (R) and 0.47 (L), and in their 60s, it was 0.56 (R) and 0.66 (L). For women in their 50s, CIMT was 0.4 (R) and 0.5 (L), and in their 60s, it was 0.6 (R) and 0.65 (L). In a report from China that examined the relationship between cardiovascular-related deaths and CIMT in four age groups (35-44, 45-54, 55-64, 65-75 years) between 2014 and 2020, the average CIMT for those aged 35-44 was 0.65 ± 0.13 mm, while for those aged 65-75, it was 0.83 ± 0.17 mm.^25^ To further investigate the relationship between our study and diseases in the elderly, it would be necessary to follow the life and medical histories of the participants. Simultaneously, other clinical indicators related to atherosclerosis, such as blood tests for cholesterol, triglycerides, and glucose, as well as dietary habits strongly associated with these parameters, should be investigated in a larger number of cases.

In conclusion, IMT showed a significant age-related increase in both men and women in various parts of the carotid artery, except for women in the bifurcation area in our study. The overall max IMT of the carotid artery showed a significant age-related increase in both men and women. While CIMT did not change much until the 40s, there was a trend of thickening in the 50s to 60s. Plaque formation began to appear around the 40s. Age-related changes in vascular diameter were more pronounced in older men, particularly those aged 50 and above. We hope that the results of this study will contribute to the prevention of strokes in the elderly.

Disclosure of Conflicts of Interest (COI): The authors report no conflicts of interest.

## Data Availability

All data produced in the present work are contained in the manuscript

https://www.jsum.or.jp/committee/diagnostic/pdf/jsum0515_guideline

